# Prospective assessment of catheter-associated bacteriuria in nursing home residents: clinical presentation, epidemiology, and colonization dynamics

**DOI:** 10.1101/2020.09.29.20204107

**Authors:** Chelsie E. Armbruster, Aimee L. Brauer, Monica S. Humby, Jiahui Shao, Saptarshi Chakraborty

## Abstract

**Background:** Catheterization facilitates continuous bacteriuria, for which the clinical significance remains unclear. This study aimed to determine the clinical presentation, epidemiology, and dynamics of bacteriuria in a cohort of long-term catheterized nursing home residents.

**Methods:** Prospective urine culture, urinalysis, chart review, and assessment of signs and symptoms of infection were performed weekly for 19 study participants over 7 months. All bacteria ≥10^3^ cfu/ml were cultured, isolated, identified, and tested for susceptibility to select antimicrobials.

**Results:** 226 of the 234 urines were polymicrobial (97%), with an average of 4.7 isolates per weekly specimen. 228 urines (97%) exhibited ≥10^6^ CFU/ml, 220 (94%) exhibited abnormal urinalysis, 126 (54%) were associated with at least one possible sign or symptom of infection, 82 (35%) would potentially meet a standardized definition of CAUTI, but only 3 had a caregiver diagnosis of CAUTI. 286 (30%) of bacterial isolates were resistant to a tested antimicrobial agent, and bacteriuria composition was remarkably stable despite a combined total of 54 catheter changes and 23 weeks of antimicrobial use.

**Conclusions:** Bacteriuria composition was largely polymicrobial, including persistent colonization by organisms previously considered to be urine culture contaminants. Neither antimicrobial use nor catheter changes sterilized the urine, at most resulting in transient reductions in bacterial burden followed by new acquisition of resistant isolates. Thus, this patient population exhibits a high prevalence of bacteriuria coupled with potential indicators of infection, necessitating further exploration to identify sensitive markers of true infection.

**Funding:** This work was supported by the NIH (R00 DK105205, R01 DK123158, UL1 TR001412)

## Introduction

Urinary catheter placement in healthcare settings is a common medical procedure utilized in the treatment of 60% of critically ill patients, 20% of those in medical and surgical intensive care units, and 5-22% of residents in long-term care facilities (1–6). This is particularly true in nursing homes, where 12-15% of newly-admitted individuals have an indwelling urinary catheter and 5-10% will have long-term urinary catheter use for indications such as chronic pressure ulcers or wounds, traumatic pelvic injury, neurogenic bladder, and having low overall functional status (4, 7–9). However, long-term urinary catheter use increases the risk of developing symptomatic catheter-associated urinary tract infection (CAUTI) and provides a reservoir for antimicrobial resistant bacteria (8, 10, 11).

Urinary catheters facilitate the presence of bacteria in urine (bacteriuria) as they bypass many of the natural defenses of the urinary tract. Bacteria that colonize the periurethral area are typically excluded from the urinary tract by a combination of micturition (the regular passing of urine), the physical barrier provided by intact urothelial cells and mucin, and by innate immune defenses. However, insertion of a catheter damages the urothelial barrier, disrupts normal micturition, and causes retention of a low volume of urine within the bladder, all of which facilitate bacterial growth (12–14). The immune response elicited by the catheter also results in accumulation of host proteins such as fibrinogen, which prime the catheter surface and facilitate bacterial attachment (15–17). The combination of these factors creates a uniquely permissive environment for numerous bacterial species to colonize and potentially establish infection. Indeed, for each day that a urinary catheter is in place, there is a 3-8% incidence of bacteriuria, and long-term catheterization (>28 days) typically results in continuous bacteriuria (1, 18).

Catheterization and resulting bacteriuria are associated with numerous adverse outcomes, such as functional decline, increased hospital stays, inappropriate or inadequate antimicrobial treatment, and an overall increase in mortality rate compared to non-catheterized individuals (1, 6, 18–23). However, catheter-associated bacteriuria is frequently asymptomatic and can be challenging to distinguish from CAUTI, especially in long-term care settings. Guidelines for diagnosis of CAUTI include the presence of clinical signs and symptoms in addition to a positive urine culture (24–26). For instance, the 2010 clinical care guidelines of the Infectious Diseases Society of America (IDSA) define CAUTI as a positive urine culture (≥10^3^ CFU/ml of ≥ 1 bacterial species) combined with signs or symptoms compatible with UTI (fever, rigors, altered mental status, malaise or lethargy with no other identified cause, flank pain, costovertebral tenderness, acute hematuria, or pelvic discomfort) in the absence of an alternate source of infection (1, 27). In the absence of these symptoms, a culture with ≥10^5^ CFU/ml of ≥1 bacterial species is considered asymptomatic catheter-associated bacteriuria (1). In contrast, the National Health Safety Network (NHSN) surveillance criteria for long-term care facilities define symptomatic CAUTI as having a positive urine culture (no more than 2 species, of which at least one must be a bacterium at ≥10^5^ CFU/ml of urine) combined with at least one of the following: a) fever (single temperature >100°F, repeated temperatures >99°F, or an increase of >2°F over baseline), b) rigors, c) new onset hypotension (<90 systolic or <60 diastolic blood pressure) without an alternate non-infectious cause, d) new onset decline in mental or functional status combined with leukocytosis (>14,000 leukocytes/mm^3^) and without an alternate diagnosis, e) new or increased suprapubic tenderness, f) new or increased costovertebral pain or tenderness, h) acute pain, swelling, or tenderness of the testes, epididymis, or prostate, or h) purulent discharge from the catheter insertion site (25, 28).

Diagnosis of CAUTI is particularly challenging in older adults, especially those with neurogenic bladder, cognitive impairments, or a high degree of functional dependence. In this population, many signs and symptoms of infection (such as fever and leukocytosis) are infrequent or absent, while others may be subtle or non-specific (29–38). A further confounding issue is the high prevalence of acute mental status change and confusion in this population (31, 36, 39), coupled with issues regarding reliable assessment of mental status and non-specific symptoms (40). To reduce inappropriate antimicrobial treatment, development of antimicrobial resistance, and risk of *Clostridioides difficile* infection, the 2019 guidelines of the IDSA strongly recommend against screening for or treating asymptomatic bacteriuria in older, functionally or cognitively impaired adults, especially those residing in long-term care facilities, as well as individuals with indwelling urinary catheters (27). The guidelines further indicate that mental status change and bacteriuria without local genitourinary symptoms or systemic symptoms should not be considered to indicate symptomatic UTI in older, functionally or cognitively impaired adults (27). However, non-specific symptoms, such as change in mental status and confusion, are the most common indications for suspected UTI in nursing home residents and antimicrobial prescription for asymptomatic bacteriuria remains common in this population (31, 41).

While several studies have reported the epidemiology of bacteriuria in catheterized individuals (1, 2, 31, 42–54), few have conducted a longitudinal assessment of colonization dynamics or included prospective assessment of common indicators of CAUTI. These collective studies have also demonstrated that polymicrobial bacteriuria and CAUTI are common during long-term catheterization, yet few report the full etiology of polymicrobial urine cultures. To address these gaps in knowledge, we conducted a prospective longitudinal assessment of bacteriuria in long-term catheterized nursing home residents from two facilities in western New York. Our study had three primary goals: 1) determine the clinical presentation of catheter-associated bacteriuria and CAUTI in long-term catheterized nursing home residents, 2) determine the epidemiology of catheter-associated bacteriuria and CAUTI, including carriage of antimicrobial resistant bacteria, and 3) determine the impact of disruptions, such as catheter changes and antimicrobial treatment, on colonization dynamics.

## Results

### Description of Study Population

Target enrollment was 50 nursing home residents with long-term indwelling urinary catheters (≥1 year), but the study was ended early due to COVID-19 with 19 participants followed for a maximum of 7 months each. As summarized in Table 1, the majority of study participants were white (79%), male (79%), and had suprapubic catheters (68%). Study participants exhibited a high level of functional dependence for activities of daily living, with an average physical self-maintenance score (PSMS) of 22 on a scale ranging from 6-30 (55). The most common comorbidities were neurogenic bladder (74%), hemiplegia (42%), diabetes (32%), renal disease (32%), multiple sclerosis (26%), and chronic heart failure (26%).

**Table 1.**
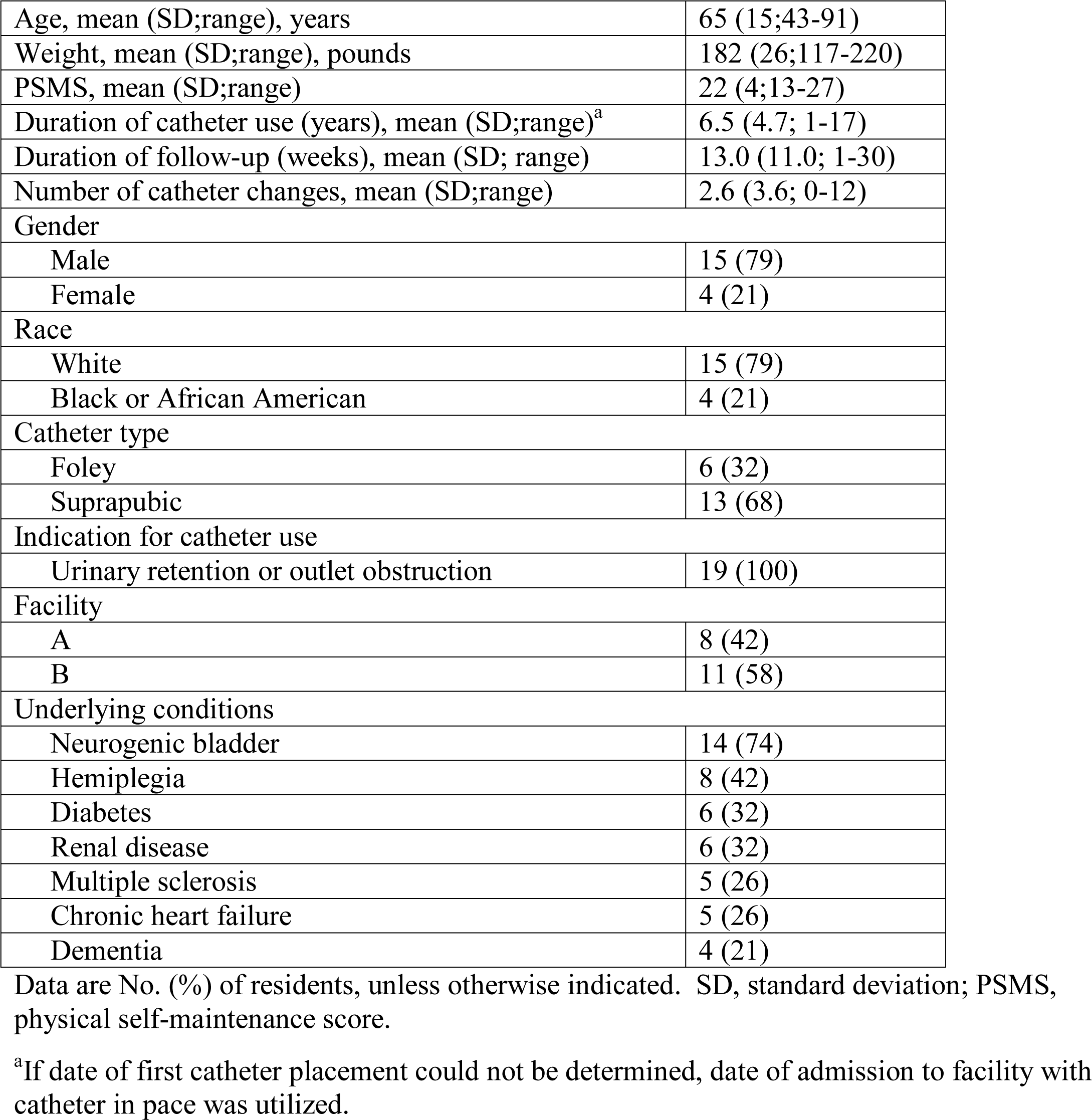
Characteristics of study participants.

|  |  |
| --- | --- |
| Age, mean (SD;range), years | 65 (15;43-91) |
| Weight, mean (SD;range), pounds | 182 (26;117-220) |
| PSMS, mean (SD;range) | 22 (4;13-27) |
| Duration of catheter use (years), mean (SD;range)<sup>a</sup> | 6.5 (4.7; 1-17) |
| Duration of follow-up (weeks), mean (SD; range) | 13.0 (11.0; 1-30) |
| Number of catheter changes, mean (SD;range) | 2.6 (3.6; 0-12) |
| Gender |  |
| Male | 15 (79) |
| Female | 4 (21) |
| Race |  |
| White | 15 (79) |
| Black or African American | 4 (21) |
| Catheter type |  |
| Foley | 6 (32) |
| Suprapubic | 13 (68) |
| Indication for catheter use |  |
| Urinary retention or outlet obstruction | 19 (100) |
| Facility |  |
| A | 8 (42) |
| B | 11 (58) |
| Underlying conditions |  |
| Neurogenic bladder | 14 (74) |
| Hemiplegia | 8 (42) |
| Diabetes | 6 (32) |
| Renal disease | 6 (32) |
| Multiple sclerosis | 5 (26) |
| Chronic heart failure | 5 (26) |
| Dementia | 4 (21) |

Data are No. (%) of residents, unless otherwise indicated. SD, standard deviation; PSMS, physical self-maintenance score.
<sup>a</sup>If date of first catheter placement could not be determined, date of admission to facility with catheter in place was utilized.

Study participants were followed for a total of 260 patient-weeks (13 average, range 1-30), with 9 of the 19 participants completing greater than 12 weeks of follow-up. Including baseline samples, 234 urine cultures were collected during the study (average 12 per participant, range 1-28). Ten participants (53%) had at least one catheter change between weekly follow-up visits, for a combined total of 54 catheter changes (5 average, range 1-12). Notably, 13 catheter changes occurred due to blockage/obstruction, 5 occurred due to accidental removal/dislodging, and the remaining 36 catheter changes were listed as routine care. Four participants (21%) experienced a combined total of 8 caregiver-diagnosed infections during follow-up, 3 of which were CAUTIs. Three participants (16%) had received antibiotics within 30 days prior to their baseline visit, and six participants (32%) received antimicrobial treatment during the course of follow-up for a total of 23 patient-weeks of antimicrobial use, including 2 participants who received antimicrobial treatment for 3 CAUTIs.

### Epidemiology of Catheter-Associated Bacteriuria

234 urine samples were collected during 260 study visits. Samples could not be obtained at 26 study visits due to absence of the participant on a particular visit or the participant not wanting to be disturbed that day. All urine samples were subjected to semi-quantitative streak plating for isolation of all distinct colony types and biochemical determination to the species level (see Methods), as well as quantitative dilution plating on HardyCHROM UTI agar to determine total colony forming units (CFU) per milliliter of urine. Of the 234 urine cultures, 233 (99%) exhibited bacterial growth >10^3^ CFU/ml, with an average of 5.12x10^6^ CFU/ml (range 5.5x10^4^ – 6.2x10^6^ CFU/ml). Notably, the only culture-negative urine was obtained from a study participant receiving intravenous antimicrobials for CAUTI with bacteremia.

A total of 1,092 bacterial isolates were cultured from 233 urines, of which 623 (57%) were Gram-negative and 469 (43%) were Gram-positive. Select antimicrobial susceptibility was assessed by zone of growth inhibition on Meuller-Hinton agar. All 623 Gram-negative isolates were tested for susceptibility to ciprofloxacin, ceftazidime, ceftazidime with clavulanate, and imipenem, all 168 *S. aureus* isolates were tested for methicillin susceptibility, and all 163 *E. faecalis* isolates were tested for vancomycin susceptibility. Of these combined 954 isolates, 286 (30%) were resistant to at least one antimicrobial agent and resistant organisms were present in urine specimens from 12 of the 19 participants (63%, Table 2). The most common resistances were ciprofloxacin (171/623 Gram-negative isolates [27%]), methicillin (81/168 *Staphylococcus aureus* isolates [48%]), and ceftazidime (37/623 Gram-negative isolates [6%]). Notably, all isolates that were resistant to ceftazidime were also resistant to ceftazidime with clavulanic acid, indicating production of an extended spectrum *β*-lactamase (ESBL). None of the Gram-negative isolates were resistant to imipenem, and none of the *E. faecalis* isolates were resistant to vancomycin.

**Table 2.** Prevalence of antimicrobial resistant isolates

| Organism | Resistance | <sup>a</sup>Participant Prevalence | <sup>b</sup>Organism Prevalence |
| --- | --- | --- | --- |
| *Staphylococcus aureus* (13 participants, 168 isolates) | Methicillin | 9 (69) | 81 (48) |
| *Proteus mirabilis* (11 participants, 118 isolates) | Ciprofloxacin | 7 (64) | 81 (69) |
| *Providencia stuartii* (9 participants, 111 isolates) | Ciprofloxacin | 4 (44) | 27 (24) |
|  | ESBL | 1 (11) | 22 (20) |
| *Morganella morganii* (5 participants, 64 isolates) | Ciprofloxacin | 2 (40) | 24 (37) |
| *Escherichia coli* (8 participants, 89 isolates) | Ciprofloxacin | 1 (12) | 26 (29) |
| *Pseudomonas aeruginosa* (8 participants, 66 isolates) | Ciprofloxacin | 3 (37) | 10 (15) |
|  | ESBL | 2 (25) | 12 (18) |
| *Proteus vulgaris* (3 participants, 37 isolates) | Ciprofloxacin | 2 (67) | 3 (8) |
| *Klebsiella oxytoca* (3 participants, 16 isolates) | ESBL | 1 (33) | 2 (12) |
| *Enterobacter aerogenes* (2 participants, 8 isolates) | ESBL | 1 (50) | 1 (12) |

<sup>a</sup>Number (percent) of study participants colonized by the organism with isolates resistant to the listed antimicrobial
<sup>b</sup>Number (percent) of isolates of the organism that were resistant to the listed antimicrobial

Antimicrobial resistance was most prevalent for *Proteus mirabilis* (69% of isolates), *S. aureus* (48%), *Providencia stuartii* (44%), *Morganella morganii* (37%), *Pseudomonas aeruginosa* (29%), and *Escherichia coli* (29%) (Table 2). For most organisms, all sequential isolates from a single participant exhibited the same colony morphology and resistance profile, with the exception of *S. aureus.* Methicillin-resistant *S. aureus* (MRSA) is therefore discussed separately from methicillin-sensitive *S. aureus* (MSSA).

The vast majority of culture-positive urines were polymicrobial (226/233, 97%), with an average of 4.7 isolates per weekly urine specimen (range 1-10); 30 (13%) harbored two distinct isolates, 24 (11%) had three isolates, 39 (17%) had four isolates, 52 (23%) had five isolates, 41 (18%) had six isolates, 18 (8%) had seven isolates, and 22 (9%) had eight or more distinct isolates. A visualization of the full colonization data for each participant at each weekly visit is provided in Supplemental Figure 1, and longitudinal colonization data from four study participants is displayed in Figure 1 to align bacteriuria composition with urinalysis and sign and symptom data. As displayed in Table 3, the most common organisms at baseline were *Enterococcus faecalis* (14/19 baseline urine specimens, 74%), *P. stuartii* (8/19, 42%), *E. coli* (7/19, 37%), coagulase-negative *Staphylococcus* (7/19, 37%), and *P. mirabilis* 6/19, 32%). When examined across all study visits, 18/19 participants (95%) were colonized by *E. faecalis* during at least one study visit, 11/19 (58%) *P. mirabilis,* 11/19 (58%) coagulase-negative *Staphylococcus,* 9/19 (47%) *P. stuartii,* 9/19 (47%) MRSA, 9/19 (47%) MSSA, 8/19/19 (42%) *E. coli,* 8/19 (42%) *P. aeruginosa,* 5/19 (26%) *M. morganii,* and 5/19 (26%) *K. pneumoniae.* Thus, the highest weekly prevalence was observed for *E. faecalis* (63%), *P. mirabilis* (45%), *P. stuartii* (43%), and *S. aureus* (33% for methicillin-sensitive isolates and 31% for methicillin-resistant isolates). The most stable and persistent colonizers were *E. faecalis, P. mirabilis, P. stuartii,* and *E. coli,* while organisms such as *P. aeruginosa, Providencia rettgeri, Klebsiella pneumoniae,* coagulase-negative *Staphylococcus,* and *Serratia marsescens* tended to exhibit transient colonization. Consistent with the high percentage of polymicrobial urines, the majority of the study participants exhibited polymicrobial bacteriuria during at least one study visit (18/19, 95%): 15/19 (79%) exhibited polymicrobial bacteriuria at all weekly visits, 3/19 (16%) mostly had polymicrobial specimens with one or two monomicrobial samples, and one participant only exhibited monomicrobial urine samples (Figure 2).

**Figure 1.**
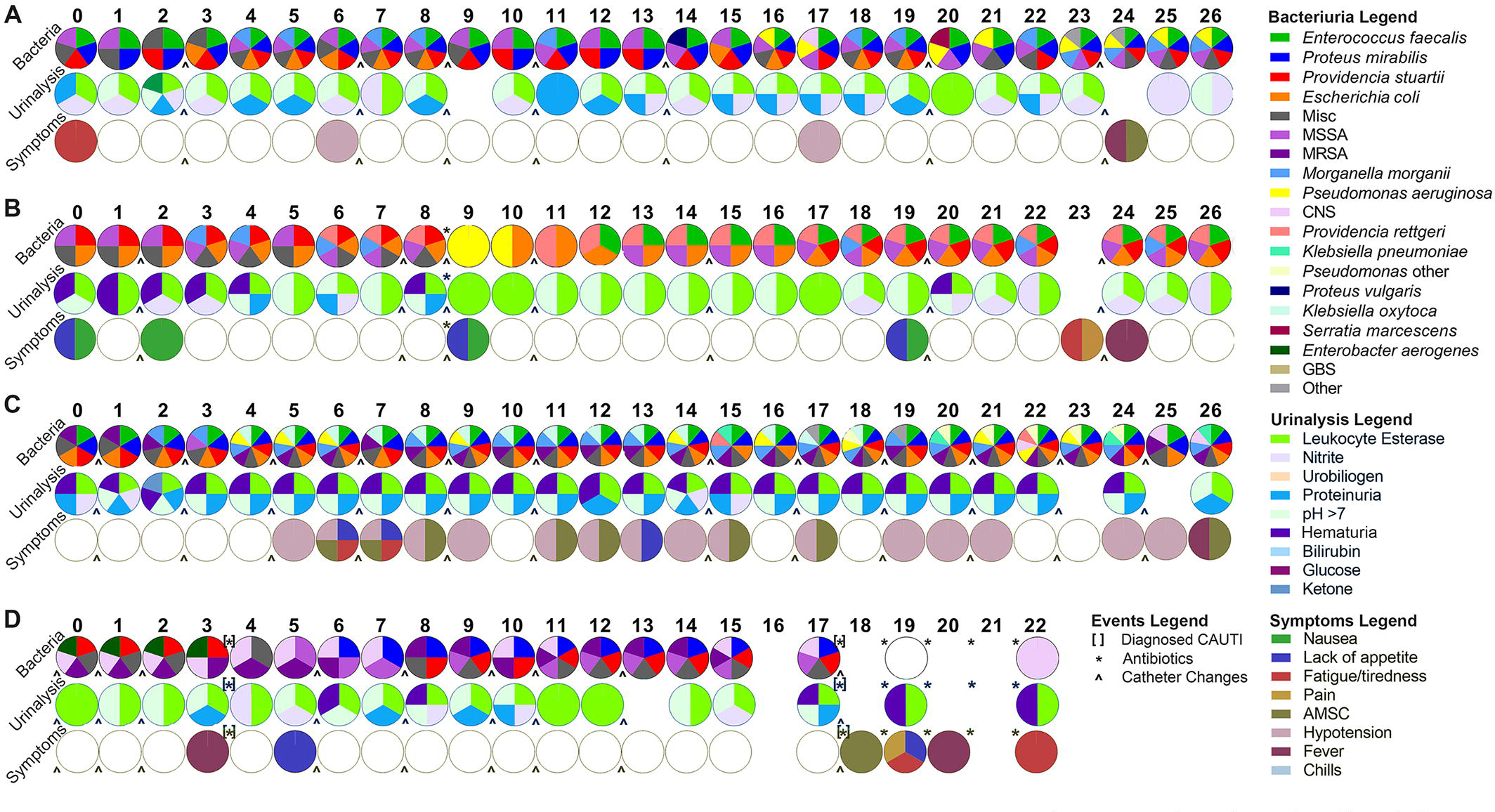
Epidemiology and dynamics of catheter-associated bacteriuria. The complete longitudinal data for four study participants (letters) at each study visit (numbers, starting with 0 for baseline) are presented. The first row of pie charts displays the bacterial species that were identified by standard culture methods at each visit, the second row displays the urinalysis test strip results, and the third row displays the potential signs and symptoms of infection that were present at each study visit. White circles with black outlines indicate study visits at which assessments were negative, while empty spaces indicate study visits at which a particular assessment could not be made. [] indicates a caregiver-diagnosed CAUTI, * indicates antimicrobial use, and ^ indicates when the participant’s catheter was changed.

**Figure 2.**
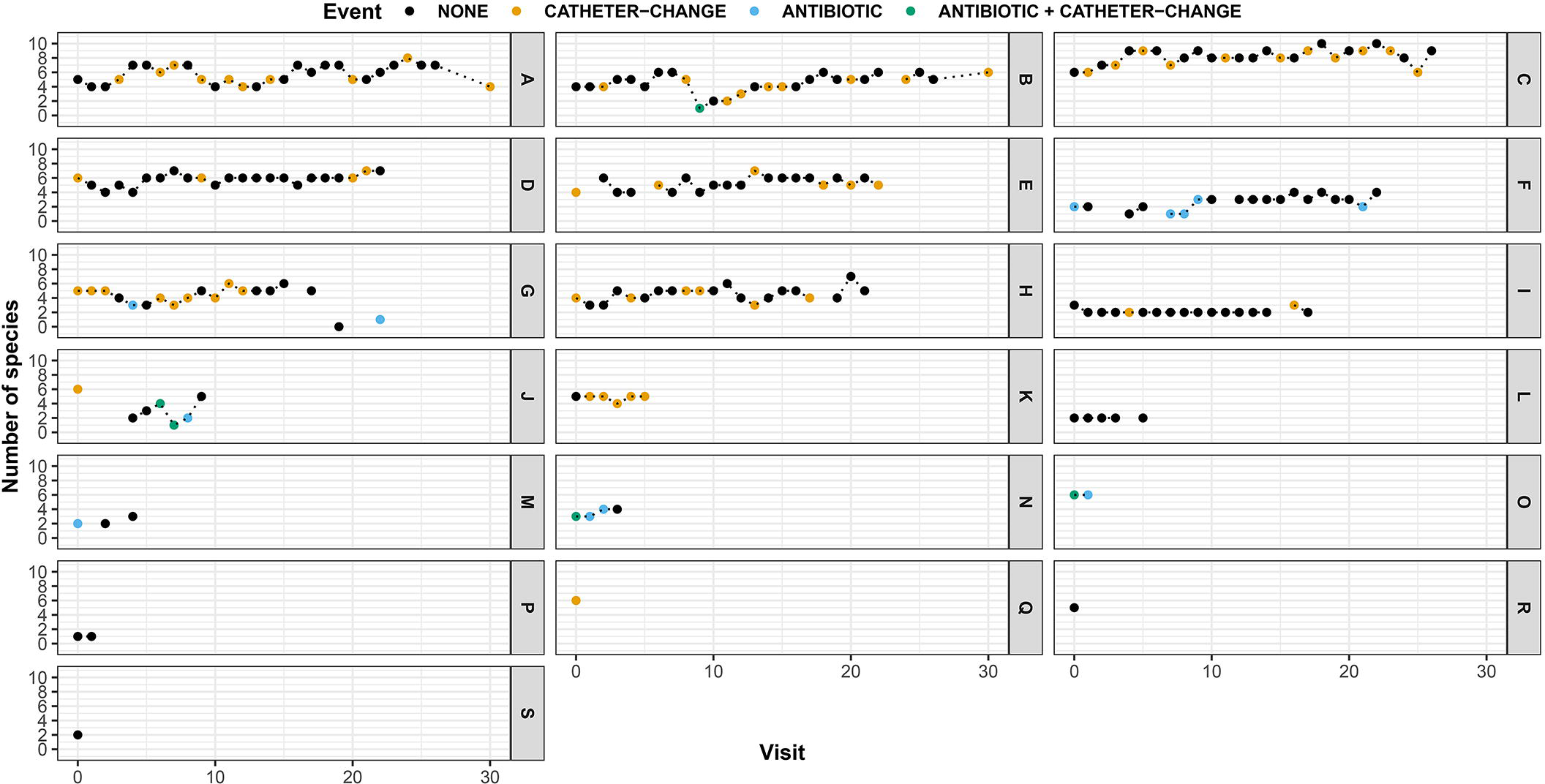
Impact of catheter changes and antibiotic use on colonization density. The total number of unique bacterial species isolated from each weekly urine sample (0-10) are displayed for all study participants (A-S) across each study visit (0-30). Symbol color indicates whether an event occurred since the prior visit that could impact colonization density; catheter change (orange), antibiotic use (light blue), or catheter change plus antibiotic use (teal).

**Table 3.**
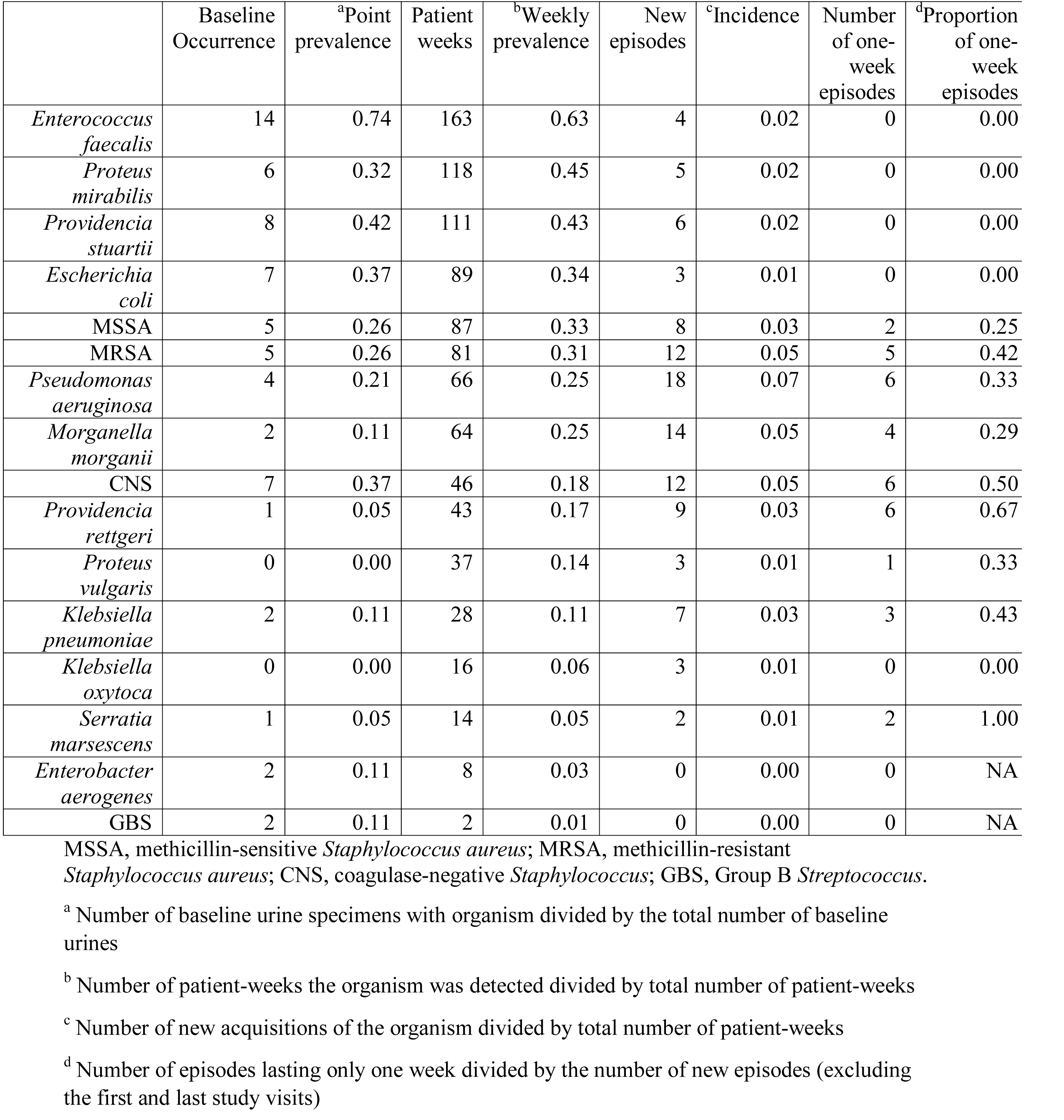
Epidemiology of Catheter-Associated Bacteriuria

|  | Baseline Occurrence | <sup>a</sup>Point prevalence | Patient weeks | <sup>b</sup>Weekly prevalence | New episodes | <sup>c</sup>Incidence | Number of one-week episodes | <sup>d</sup>Proportion of one-week episodes |
| --- | --- | --- | --- | --- | --- | --- | --- | --- |
| *Enterococcus faecalis* | 14 | 0.74 | 163 | 0.63 | 4 | 0.02 | 0 | 0.00 |
| *Proteus mirabilis* | 6 | 0.32 | 118 | 0.45 | 5 | 0.02 | 0 | 0.00 |
| *Providencia stuartii* | 8 | 0.42 | 111 | 0.43 | 6 | 0.02 | 0 | 0.00 |
| *Escherichia coli* | 7 | 0.37 | 89 | 0.34 | 3 | 0.01 | 0 | 0.00 |
| MSSA | 5 | 0.26 | 87 | 0.33 | 8 | 0.03 | 2 | 0.25 |
| MRSA | 5 | 0.26 | 81 | 0.31 | 12 | 0.05 | 5 | 0.42 |
| *Pseudomonas aeruginosa* | 4 | 0.21 | 66 | 0.25 | 18 | 0.07 | 6 | 0.33 |
| *Morganella morganii* | 2 | 0.11 | 64 | 0.25 | 14 | 0.05 | 4 | 0.29 |
| CNS | 7 | 0.37 | 46 | 0.18 | 12 | 0.05 | 6 | 0.50 |
| *Providencia rettgeri* | 1 | 0.05 | 43 | 0.17 | 9 | 0.03 | 6 | 0.67 |
| *Proteus vulgaris* | 0 | 0.00 | 37 | 0.14 | 3 | 0.01 | 1 | 0.33 |
| *Klebsiella pneumoniae* | 2 | 0.11 | 28 | 0.11 | 7 | 0.03 | 3 | 0.43 |
| *Klebsiella oxytoca* | 0 | 0.00 | 16 | 0.06 | 3 | 0.01 | 0 | 0.00 |
| *Serratia marsescens* | 1 | 0.05 | 14 | 0.05 | 2 | 0.01 | 2 | 1.00 |
| *Enterobacter aerogenes* | 2 | 0.11 | 8 | 0.03 | 0 | 0.00 | 0 | NA |
| GBS | 2 | 0.11 | 2 | 0.01 | 0 | 0.00 | 0 | NA |

MSSA, methicillin-sensitive *Staphylococcus aureus*; MRSA, methicillin-resistant *Staphylococcus aureus*; CNS, coagulase-negative *Staphylococcus*; GBS, Group B *Streptococcus*.
<sup>a</sup> Number of baseline urine specimens with organism divided by the total number of baseline urines
<sup>b</sup> Number of patient-weeks the organism was detected divided by total number of patient-weeks
<sup>c</sup> Number of new acquisitions of the organism divided by total number of patient-weeks
<sup>d</sup> Number of episodes lasting only one week divided by the number of new episodes (excluding the first and last study visits)

Microbe-microbe interactions during polymicrobial colonization can have profound implications for risk of developing severe disease (56–61). To quantify such interactions, we first computed the number of occurrences for each microbial species (i.e., the total number of participants colonized by that species) at each time point across the entire longitudinal dataset. We then computed Lin’s concordance correlation coefficient (62) between the number of occurrences of each microbial pair over the entire longitudinal dataset (Supplemental Table 1). Highly concordant co-colonization partners included *P. mirabilis* with *P. stuartii* (80/233 culture-positive urines [34%]; 8/19 participants [42%])*, P. mirabilis* with *M. morganii* (52/233 [22%]; 4/19 [21%])*, M. morganii* with *P. stuartii* (63/233 [27%]; 5/19 [26%])*, P. mirabilis* with *E. faecalis* (93/233 [40%]; 9/19 [47%]), and *P. stuartii* with *E. faecalis* (86/233 [37%]; 8/19 [42%]). A total of 205 urine samples from all 19 study participants contained at least one of these four organisms; 121 (59%) samples from 11 participants contained at least two of the four concordant organisms, 71 (35%) samples from 7 participants contained at least three, and 50 (24%) samples from 4 participants included all four organisms. It is also notable that co-colonization by these species occurred in both men and women, and with both catheter types (Foley *versus* suprapubic).

### Clinical Presentation of Catheter-Associated Bacteriuria

Potential signs and symptoms of infection were prospectively assessed by a nurse at each study visit, and also collected from participant medical records to determine point prevalence at baseline, weekly prevalence, and incidence (Table 4). Assessment included pain (defined as suprapubic and/or costovertebral pain or tenderness), fever (defined as temperature >99°F), hypotension (defined as <90 systolic or <60 diastolic blood pressure), acute mental status change (defined as altered level of consciousness, inattention, or disorganized thinking in one of two mini-cognitive assessments conducted 30 minutes apart), nausea, lack of appetite, and fatigue.

**Table 4.** Clinical Presentation of Catheter-Associated Bacteriuria

|  | Baseline Occurrence | <sup>a</sup>Point prevalence | Patient weeks | <sup>b</sup>Weekly prevalence | New episodes | <sup>c</sup>Incidence | Number of one-week episodes | <sup>d</sup>Proportion of one-week episodes |
| --- | --- | --- | --- | --- | --- | --- | --- | --- |
| ≥10<sup>5</sup> CFU/ml | 19 | 1.00 | 231 | 0.99 | 1 | 0.00 | 0 | 0.00 |
| Hypotension | 3 | 0.16 | 36 | 0.14 | 14 | 0.05 | 7 | 0.50 |
| Fatigue | 3 | 0.16 | 46 | 0.18 | 13 | 0.05 | 8 | 0.62 |
| <sup>e</sup>Altered mental status | 2 | 0.11 | 48 | 0.18 | 26 | 0.10 | 14 | 0.54 |
| Lack of appetite | 3 | 0.16 | 43 | 0.17 | 17 | 0.07 | 12 | 0.71 |
| <sup>f</sup>Pain | 2 | 0.11 | 38 | 0.15 | 10 | 0.04 | 6 | 0.60 |
| Nausea | 2 | 0.11 | 31 | 0.12 | 11 | 0.04 | 5 | 0.45 |
| <sup>g</sup>Fever | 0 | 0.00 | 11 | 0.04 | 11 | 0.04 | 11 | 1.00 |
| Chills | 0 | 0.00 | 10 | 0.04 | 7 | 0.03 | 4 | 0.57 |
| Leukocytosis | 0 | 0.00 | 2 | 0.01 | 2 | 0.01 | 2 | 1.00 |

<sup>a</sup> Number of baseline visits with symptom divided by the total number of baseline visits
<sup>b</sup> Number of patient-weeks symptom was detected divided by total number of patient-weeks
<sup>c</sup> Number of new instances of symptom divided by total number of patient-weeks
<sup>d</sup> Number of episodes lasting only one week divided by the number of new episodes (excluding the first and last study visits)
<sup>e</sup> Altered mental status was assessed by bCAM and DTS (see methods)
<sup>f</sup> Pain refers to costovertebral or suprapubic pain or tenderness
<sup>g</sup> Fever was defined as having a temperature >99°F

The sign and symptom assessment tool utilized by the study nurses is provided in Supplemental Figure 2. Overall, 16/19 (84%) participants exhibited potential signs and symptoms of infection at 126/260 (48%) study visits. A visualization of the sign and symptom data for each participant at each weekly visit is provided in Supplemental Figure 3, and data aligned to bacteriuria and urinalysis results from four participants are displayed in Figure 1.

The most common clinical presentations at baseline were hypotension (3/19, 16%), fatigue (3/19, 16%), and lack of appetite (3/19, 16%). Altered mental status, fatigue, and lack of appetite exhibited the highest weekly prevalence and incidence, while fever, chills, and leukocytosis were absent at baseline, had a low weekly prevalence and incidence, and were typically only present at a single study visit. Lin’s concordance correlation coefficients between the number of occurrences of sign and symptom pairs across the entire longitudinal dataset displayed a high degree of concordance between nausea, lack of appetite, fatigue or tiredness, and pain (Supplemental Table 2). Of the 14 participants who completed at least 4 weeks of follow-up, 3 (21%) were asymptomatic for the vast majority of visits, 3 (21%) experienced one or two symptomatic episodes that lasted approximately three weeks (including three of the four individuals who experienced caregiver-diagnosed infections), 4 (29%) experienced frequent multi-week stretches of potential signs or symptoms of infection interspersed with one or two asymptomatic weeks, and 4 (29%) exhibited potential signs or symptoms of infection at every study visit. These data highlight some of the challenges for diagnosing CAUTI in long-term catheterized nursing home residents, including the high prevalence and subjective nature of constitutional signs and symptoms of infection (such as nausea, lack of appetite, and fatigue) in this patient population.

Due to the high burden of bacteriuria and possible signs and symptoms of infection in this population, 82/260 (31%) study visits would potentially meet the IDSA CAUTI criteria (1, 27). If only new-onset symptoms are considered, this number is reduced to 51/260 (20%). Only 6 study visits would meet the more strict NHSN surveillance definition for CAUTI (25, 28) due to the stringent cutoff of ≤2 organisms in the urine culture and the requirement that acute mental status change must be accompanied by leukocytosis (which was not prospectively assessed and only reported in the medical records of 3 study participants). Importantly, none of the 3 caregiver-diagnosed CAUTIs would meet the NHSN surveillance criteria since all 3 involved >2 organisms. However, 2 of the 3 diagnosed CAUTIs involved bacteremia with one of the bacterial species present in urine (Supplemental Table 3), indicating that these cases likely represent true CAUTI. Supplemental Table 3 also demonstrates substantial concordance between the bacterial culture results and sign and symptom analysis conducted by the study team and those reported in the participants’ medical records. If urine cultures containing 3 or more organisms are permitted for the NHSN criteria, the number of visits that would meet the NHSN CAUTI surveillance definition increases to 27 and includes all 3 caregiver-diagnosed CAUTIs. Taken together, prospective weekly assessment of possible signs and symptoms of CAUTI revealed that numerous potential indicators of infection are common in nursing home residents with long-term catheters and fluctuate in duration, which may further confound the utility of these indicators for distinguishing CAUTI from asymptomatic bacteriuria in this patient population.

All urine samples were also subjected to urinalysis via 10-parameter urine reagent test strip to determine the point prevalence of parameters that are often considered to be suggestive of urinary tract infection, along with weekly prevalence and incidence (Table 5). A visualization of the urinalysis data for each participant at each weekly visit is provided in Supplemental Figure 4, and aligned with bacteriuria and symptom data for four participants in Figure 1. The most common urinalysis findings at baseline were a positive result for leukocyte esterase (17/19 participants, 89%), nitrites (10/19, 53%) hematuria (8/19, 42%), and proteinuria (5/19, 26%).

**Table 5.** Urinalysis of Catheter-Associated Bacteriuria

|  | Baseline Occurrence | <sup>a</sup>Point prevalence | Patient weeks | <sup>b</sup>Weekly prevalence | New episodes | <sup>c</sup>Incidence | Number of one-week episodes | <sup>d</sup>Proportion of one-week episodes |
| --- | --- | --- | --- | --- | --- | --- | --- | --- |
| Leukocytes | 17 | 0.89 | 213 | 0.91 | 8 | 0.03 | 0 | 0 |
| Protein | 10 | 0.53 | 131 | 0.56 | 30 | 0.13 | 13 | 0.43 |
| Nitrite | 10 | 0.53 | 124 | 0.53 | 36 | 0.15 | 16 | 0.44 |
| Blood | 8 | 0.42 | 75 | 0.32 | 14 | 0.06 | 12 | 0.86 |
| pH $\geq 7.25$ | 4 | 0.21 | 105 | 0.45 | 29 | 0.12 | 4 | 0.14 |
| Ketone | 0 | 0.00 | 14 | 0.06 | 9 | 0.04 | 6 | 0.67 |
| Urobilinogen | 0 | 0.00 | 2 | 0.01 | 2 | 0.01 | 2 | 1.00 |
| Bilirubin | 0 | 0.00 | 2 | 0.01 | 2 | 0.01 | 2 | 1.00 |
| Glucose | 0 | 0.00 | 0 | 0.00 | 0 | 0.00 | 0 | 0.00 |

<sup>a</sup> Number of baseline urine urinalysis finding divided by the total number of baseline urines
<sup>b</sup> Number of patient-weeks urinalysis finding was detected divided by total number of patient- weeks
<sup>c</sup> Number of new instances of urinalysis finding divided by total number of patient-weeks
<sup>d</sup> Number of episodes lasting only one week divided by the number of new episodes (excluding the first and last study visits)

The highest weekly prevalence was observed for leukocyte esterase, proteinuria, and nitrites. Leukocyte esterase and high pH were highly persistent and typically identified in multiple consecutive urine samples, while hematuria and ketones were more likely to be present at a single study visit. Overall, 226 of the 227 urine specimens tested (99%) had a urinalysis result that could be suggestive of infection in a non-catheterized individual and may therefore influence perception of urine culture results. While this is likely just a reflection of the ubiquitous bacteriuria experienced by this patient population, these data underscore the need for caution when interpreting urinalysis results and possible signs and symptoms of urinary tract infection in nursing home residents with long-term indwelling catheters.

### Impact of Catheter Changes and Antibiotic use on Colonization Dynamics and Symptom Burden

The epidemiology of bacteriuria was remarkably consistent across consecutive study visits, often with only minor or transient perturbations. We therefore sought to determine the impact of catheter changes and antimicrobial use on colonization dynamics for all instances with at least three weeks of post-event follow-up (Table 6). Eight participants with at least 12 weeks of follow-up had an average of 5.75 catheter changes each (range 0-12). Catheter changes resulted in a combined loss of 28 isolates, with an average loss of 3.5 per participant (Table 6). However, 20 of the 28 isolates (71%) were regained within the subsequent 4 weeks, indicating that catheter changes were not effective in reducing bacteriuria long-term. Strikingly, 23 new isolates were acquired after catheter changes, for an average of 2.9 new isolates per participant gained from catheter changes. Altogether, catheter changes resulted in a net increase of 1.9 isolates, indicating that changing the catheter was more likely to introduce new organisms than reduce colonization by existing organisms. Furthermore, four participants (50%) acquired at least one new isolate resistant to a tested antimicrobial after a catheter change: 5 of the 23 new acquisitions were resistant organisms (22%), resulting in 28% average incidence of new acquisition of resistant organisms following catheter changes. Importantly, there was no common source of transmission of new resistant organisms among these four participants as two resided at facility A and two resided at facility, and all were from different units on different floors within each facility.

**Table 6.** Colonization Dynamics

| Participant | Number of events | Isolates lost<sup>a</sup> | Isolates re-gained<sup>b</sup> | New isolates<sup>c</sup> | New ARO<sup>d</sup> | Incidence of new ARO<sup>e</sup> | Net change<sup>f</sup> |
| --- | --- | --- | --- | --- | --- | --- | --- |
| Catheter Changes |  |  |  |  |  |  |  |
| A | 7 | 7 | 4 | 6 | 0 | NA | +3 |
| B | 5 | 1 | 0 | 0 | 1 | 0.17 | -1 |
| C | 11 | 12 | 11 | 9 | 1 | 0.11 | +8 |
| D | 2 | 0 | 0 | 1 | 0 | NA | +1 |
| E | 2 | 2 | 0 | 5 | 1 | 1.00 | +3 |
| G | 7 | 4 | 4 | 1 | 2 | 0.40 | +1 |
| H | 4 | 2 | 1 | 0 | 0 | 0.00 | -1 |
| I | 2 | 0 | 0 | 1 | 0 | NA | +1 |
| *Average* | 5.0 | 3.5 | 2.5 | 2.9 | 0.6 | 0.28 | +1.9 |
| Antimicrobial Treatments |  |  |  |  |  |  |  |
| B | 1 | 5 | 3 | 2 | 0 | 0.00 | 0 |
| F | 1 | 1 | 0 | 2 | 0 | 0.00 | +1 |
| G | 1 | 2 | 1 | 1 | 1 | 0.50 | 0 |
| *Average* | 1.0 | 2.7 | 1.3 | 1.7 | 0.3 | 0.17 | +0.3 |

Data pertain to all catheter changes and antimicrobial treatments with at least 3 weeks of subsequent follow-up from 9 study participants who completed at least 16 weeks of follow-up visits.
<sup>a</sup> Number of isolates present the week prior to the event and absent the week following the event
<sup>b</sup> Number of isolates that were lost due to the event but reemerged in the subsequent three weeks
<sup>c</sup> Number of isolates that were not present the week prior to the event but emerged in the subsequent three weeks
<sup>d</sup> Number of new isolates that were antimicrobial resistant organisms (AROs)
<sup>e</sup> Number of new ARO acquisitions divided by total number of new acquisitions
<sup>r</sup> Net impact of the event on the number of isolates detected in the participants weekly urine samples
NA, Not applicable

In addition to catheter changes, three participants with at least 12 weeks of follow-up each received at least one patient-week of antimicrobial treatment with at least 3 weeks of subsequent follow-up (Table 6). Treatment resulted in a combined loss of 8 isolates, with an average loss of 2.7 colonizing isolates per participant. Similar to catheter changes, 4 of the 8 isolates (50%) were regained within the subsequent 3 weeks, and 5 new isolates were acquired (average of 1.7 per participant) resulting in a net increase of 0.3 isolates after antimicrobial treatment. One of the 5 new isolates was resistant to a tested antimicrobial (20%), resulting in a 17% average incidence of new acquisition of resistant organisms following treatment.

It is also notable that 15/27 (56%) study visits during which the NHSN CAUTI criteria would be met if bacteriuria with ≥3 organisms is permitted occurred within 10 days after a catheter change or antibiotic use, including all 3 of the caregiver-diagnosed CAUTIs. To further explore the impact of catheter changes and antibiotic use on colonization dynamics and symptom burden, two multivariate Bayesian multilevel models were used to analyze the entire dataset to determine the contribution of parameters at any given visit to the likelihood of a specific microbe (Supplemental Figure 5) or symptom (Supplemental Figure 6) being present at the following study visit. The Bayesian models coherently address missingness in the data, and provide probabilistic quantification of modeling uncertainties (see Methods for details). For bacterial colonization, the main contributor to colonization by a specific microbe at any given study visit was the presence of that same microbe at the prior study visit, which reflects the overall high degree of stability in colonization. Interestingly, neither antibiotic use nor catheter changes significantly altered the likelihood of colonization by any specific microbe in this analysis, although antibiotic use resulted in a trend towards decreased likelihood of colonization by MSSA, *P. rettgeri, P. stuartii, M. morganii, P. mirabilis, E. faecalis,* and miscellaneous PYR-negative Catalase-negative Gram-positive isolates as well as a trend towards increased likelihood of colonization by *P. aeruginosa* and coagulase-negative *Staphylococcus* species. In agreement with our qualitative assessment of the impact of catheter changes and antibiotic treatment on colonization over time (Table 6), neither event significantly reduced the total number of bacterial species present in urine samples from week to week (95% posterior credible intervals for the odds ratios are 0.9994-1.0022 and 0.5546-1.0003 respectively, Figure 2). Similar results were observed for analysis of possible signs and symptoms of infection, with the main contributor to the presence of most symptoms at any given study visit being the presence of that same symptom at the prior study visit, especially for hypotension, pain, fatigue or tiredness, and acute mental status change. Antibiotic use resulted in a slight trend towards increased likelihood of nausea and chills, while catheter changes had no apparent impact in this analysis.

## Discussion

CAUTIs are common in nursing home residents with long-term indwelling catheters and the leading cause of antimicrobial prescriptions in this population (63). However, it is estimated that approximately one-third of CAUTIs are misdiagnosed asymptomatic bacteriuria, for which antimicrobial therapy is not considered to be beneficial (64, 65). Part of the discrepancy is due to the challenges of diagnosing CAUTI, especially in a patient population that frequently presents with atypical symptoms. While our sample size was small, our study clearly demonstrates that nursing home residents with long-term catheters routinely have bacteriuria >10^5^ CFU/ml combined with abnormal urinalysis results and numerous possible signs and symptoms of urinary tract infection, which underscores the difficulties of diagnosing true infection in this population.

Interestingly, there were no clear differences in the composition of bacteriuria between weekly asymptomatic cultures, study visits that would meet either the IDSA or NHSN CAUTI criteria, and study visits at which participants had a caregiver diagnosis of CAUTI. It is particularly striking that the organisms present at the time of all three caregiver diagnosed CAUTIs had been persisting in the participants’ urine specimens for several weeks prior to symptom onset and diagnosis (see participants G and J in Supplemental Figure 1). Further investigation of urine specimens using metagenomics and proteomics will be necessary to determine if development of signs and symptoms of infection correlates with the presence of specific organisms including those not detected by standard culture techniques such as fungi, anaerobic bacteria, and viruses, or to changes in the host immune response. Assessment of a larger patient population will also be necessary to determine which combinations of factors provide the greatest sensitivity and specificity for CAUTI requiring therapeutic intervention *versus* asymptomatic colonization.

Over-testing of urine samples, both by culture and urinalysis, has been demonstrated to have a predominantly negative impact on patient outcomes including inappropriate antimicrobial prescription and increased duration of hospitalization (66–72). For instance, detection of pyuria in preoperative urinalysis was recently demonstrated be associated with prescription of antimicrobials even in the absence of a positive urine culture or urinary symptoms (73). Importantly, pyuria itself was not associated with any negative postoperative outcomes, but the resulting antimicrobial use increased risk of subsequent *Clostridioides difficile* infection without improving any other outcomes (73). This issue is further complicated in catheterized individuals, older adults, and those with neurogenic lower urinary tract dysfunction, for whom pyuria and positive urine cultures are common, and fever, dysuria, urgency, and pain demonstrate minimal sensitivity and specificity for differentiating UTI from asymptomatic bacteriuria (27, 68, 74–79). A recent study identified fever as the primary indication for obtaining a urine culture from catheterized individuals, even when other urinary symptoms were lacking and there were possible alternative explanations of fever (80). Abnormal urinalysis is another common indication for obtaining a urine culture in hospital settings (68), despite lack of other urinary symptoms and IDSA guidelines to the contrary. Considering the almost ubiquitous bacteriuria and abnormal urinalysis results observed in this study, coupled with a high prevalence of non-specific signs and symptoms of possible infection, our data underscore the critical need for discovery of additional indicators of true infection in this patient population.

Frequent exposure to antimicrobials has been demonstrated to result in a high carriage rate of antimicrobial resistant organisms in nursing home residents, particularly those with indwelling devices (8, 10, 11). While only 6 of the 19 participants received antimicrobials during the course of the study, 12 participants (63%) were colonized by at least one bacterium that was resistant to a tested antimicrobial, and 26% of the 234 urine cultures contained at least one resistant bacterium. The persistence of antimicrobial resistant isolates was also striking. For instance, of the 9 participants with MRSA bacteriuria, 4 exhibited MRSA for at least 8 weeks despite multiple catheter changes, and similar trends were observed for resistant Gram-negative bacteria.

Regarding persistent colonization, our data indicate that bacteriuria remains remarkably stable in individuals with long-term urinary catheters even after numerous catheter changes, and antimicrobial treatment only transiently reduced colonization of some of the organisms. It is alarming that antibiotic use did not sterilize the urine for any of the study participants, with the exception of a single urine sample taken from participant G while they were undergoing several weeks of intravenous antibiotic exposure. While antibiotic use often resulted in an immediate loss of one or two colonizing organisms, the net change in colonization favored an eventual increase back to the same total number of species present prior to treatment, and often included new acquisition of a resistant bacterium. Changing the catheter upon initiation of antimicrobial treatment has been suggested to expand the duration of post-treatment culture-negative bacteriuria (81), but there is no clear improvement of clinical outcomes (82). Catheter insertions can also have a substantial negative impact due to the risk of creating a false passage, bladder perforation, external trauma, and hematuria, in addition to potentially inducing symptomatic CAUTI (71). Thus, our exploratory data suggest that catheter changes in long-term catheterized nursing home residents with asymptomatic bacteriuria may impose a risk of increased acquisition of bacteria as well as new onset of signs and symptoms that are often considered to be indicators of infection without substantially reducing bacterial burden. It is important to note that not all of the reported catheter changes could have been avoided, as 13 of the 54 changes (24%) were due to catheter obstruction or blockage and 5 (9%) were due to accidental removal or dislodging of the catheter. However, the remaining 36 (67%) were reported as routine care and could potentially have been avoided. While further studies are necessary, these preliminary observations provide support for catheter care practices of only changing the catheter when necessary and strictly adhering to stewardship guidelines.

The prevalence of polymicrobial bacteriuria during long-term catheterization has been widely reported for decades (1, 2, 50). However, polymicrobial clinical urine specimens are often suspected of harboring periurethral or vaginal microbiota, particularly when they include Gram-positive organisms (83, 84). This has complicated investigation of the clinical significance of polymicrobial bacteriuria and assessment of the contribution of these organisms to pathogenesis. It is therefore notable that prospective assessment of bacteriuria revealed that *Enterococcus faecalis, Staphylococcus aureus,* and coagulase-negative *Staphylococcus* are prevalent and persistent constituents of bacteriuria in nursing home residents with long-term indwelling catheters, especially as they may facilitate transient bacteremia, hematogenous seeding of other body sites, and endocarditis (85). In elderly catheterized individuals, ∼4% of catheter changes were demonstrated to result in transient bacteremia, including by coagulase-negative *Staphylococcus* species (1, 86). Bacteremia due to *S. aureus* has also been observed in ∼7% of patients with *S. aureus* bacteriuria, particularly those of advanced age or residing in nursing homes (87). Furthermore, identification of *S. aureus* bacteriuria ≥48 hours prior to bacteremia was associated with an increased risk of mortality (87). Further research is necessary to determine if persistent catheter-associated bacteriuria with Gram-positive organisms increases risk of bacteremia and hematogenous seeding of other body sites, and if polymicrobial bacteriuria further modifies risk.

Overall, the most frequent and persistent cause of polymicrobial bacteriuria in this study was *E. faecalis* with *P. mirabilis.* The association of these organisms may have important clinical implications, as interactions between *P. mirabilis* and *E. faecalis* increase the likelihood of developing urinary stones and bacteremia in experimental models of CAUTI (17, 60). Ineffective antimicrobial treatment has also been reported to be more common for polymicrobial UTI and those involving *E. faecalis* (23). It may therefore be hypothesized that CAUTI sequelae and mortality may be more common in co-colonized study participants than those who were not co-colonized. While the present study was not sufficiently powered to address this hypothesis, it is worth noting that three study participants developed urinary stones, pyelonephritis, or urosepsis (rows D, G, and J of Supplemental Figure 1); all three were colonized by *P. mirabilis,* and two exhibited co-colonization with *E. faecalis.* All three participants were also co-colonized by *Providencia stuartii,* which has similarly been shown to interact with *P. mirabilis* and enhance risk of urinary stones and bacteremia (17, 88). However, other co-colonized participants did not exhibit infection or sequelae (such as rows A, C, and E), underscoring that these polymicrobial interactions are one of many factors that contribute to the risk of developing severe disease. Further investigation of complex polymicrobial interactions in the catheterized urinary tract are likely to provide new insight into potential decolonization strategies or therapeutics to reduce the risk of progressing from asymptomatic bacteriuria to CAUTI and associated sequelae.

The results of this study should be considered in light of several strengths and weaknesses. Main strengths of the study include 1) prospective longitudinal urine culturing rather than collection of a single specimen per participant; 2) prospective assessment of possible signs and symptoms of infection; 3) weekly study visits conducted by the same study personnel; 4) weekly urine culturing and antimicrobial susceptibility testing to identify and characterize all isolates prior to any laboratory adaptation and to monitor dynamics of colonization; and 5) enrollment at two nursing facilities. Limitations of the study include the limited sample size and exploratory nature of the study, lack of a control group for assessment of the impact of catheter changes and antibiotic use, low diversity of study participants, limited duration of follow-up for some study participants, inability to conduct assessments for all participants at all study visits, and use of biochemical tests to identify bacterial isolates rather than a more sensitive technology such as matrix assisted laser desorption ionization-time of flight mass spectrometry (MALSI-TOF MS). While our study is exploratory in nature and these limitations preclude more sophisticated analysis of the relationship between catheter changes, antibiotic use, presence of specific organisms, and clinical presentation, this study still represents a significant advance in our understanding of the dynamics and epidemiology of bacteriuria in nursing home residents with long-term urinary catheters. Further investigations of this nature may reveal the host and microbial factors that provide the greatest sensitivity and specificity for CAUTI requiring therapeutic intervention *versus* asymptomatic colonization. If so, this information could help in refining existing tools and determining which course of action should be taken for a given patient, therefore guiding appropriate antimicrobial treatment and possibly reducing acquisition of antimicrobial resistance.

## Methods

### Ethics statement

This study was approved by the University at Buffalo Institutional Review Board (STUDY00002526) and complied with the provisions of the Declaration of Helsinki, Good Clinical Practice guidelines, and local laws and regulations. All participants (or approved decision makers) provided written informed consent prior to initiation of investigation, and all participants also assented to being in the study.

### Study design

A prospective observational cohort study of asymptomatic catheter-associated bacteriuria was conducted at two nursing homes located in Buffalo, New York between July 2019 and March 2020. Study visits occurred at enrollment and weekly thereafter for up to 7 months. Each study visit entailed chart review by trained research staff as well as a brief assessment of possible signs and symptoms of infection and collection of a urine specimen by one of three licensed practicing nurses (LPNs) from the Visiting Nurse Association of Western New York. Participants were withdrawn from the study upon indication that they no longer wanted to participate, removal of the indwelling catheter without replacement, transfer to a non-participating facility, or death. All study data and records were managed using REDCap (Research Electronic Data Capture) tools (89, 90), hosted through the University at Buffalo Clinical and Translational Science Institute.

### Inclusion criteria

Nursing home residents at either of the two participating facilities were eligible for inclusion if they had an indwelling urinary catheter (Foley or suprapubic) for at least 12 months, were at least 21 years of age, were capable of assenting to participation, and informed consent could be obtained from the resident or approved decision maker. Residents receiving end-of-life care were excluded from the study.

### Data collection from chart review

Information pertaining to participant demographics, age, weight, gender, comorbidities, functional status, indication for catheterization, duration of indwelling catheter use, history of urinary tract infection, and history of antimicrobial use were obtained from participant medical records by trained research staff on the baseline visit. Chart reviews were also conducted at each weekly study visit to obtain information pertaining suspected infections, hospitalizations, urine culture results, urinalysis results, and antimicrobial prescriptions. Potential signs and symptoms of CAUTI that were recorded include fever (defined as having a single temperature >100°F or repeated temperatures >99°F or >2°F above baseline), suprapubic or costovertebral pain or tenderness, hypotension, chills or rigors, and acute mental status change (defined as a fluctuation in behavior, inattention, disorganized thinking, or an altered level of consciousness compared to baseline) (24, 25).

If a study participant was temporarily transferred to a hospital, medical records from the hospital stay were utilized to obtain information pertaining to suspected infections.

### Assessment of possible signs and symptoms of infection

At each study visit, an LPN collected vital signs (tympanic temperature and blood pressure) and assessed costovertebral and suprapubic pain or tenderness. The LPN and a study team member also conducted a Delirium Triage Screen (DTS) (91) at the start of each visit and a Brief Confusion Assessment Method (bCAM) (91) assessment at the end of each visit to identify altered mental status (defined as fluctuating altered mental status, including altered level of consciousness, inattention, and disorganized thinking). The LPN and study team member also administered an oral questionnaire at each study visit to determine if the participant had experienced rigors or chills, nausea, lack of appetite, or fatigue since the previous visit. The sign and symptom assessment tool utilized by the study nurses is provided in Supplemental Figure 2.

### Urine collection

Urine specimens were collected from the port of the indwelling catheter by an LPN using aseptic technique. Briefly, the catheter tubing was clamped ∼12 inches below the latex rubber port and urine was allowed to collect for approximately 30 minutes. The catheter port was then swabbed with an alcohol wipe and allowed to dry for ∼30 seconds. The needle of a sterile syringe was then inserted into the port, and urine was withdrawn and transferred into a sterile specimen jar. Urine specimens were placed in an insulated cooler with ice packs and stored therein for no more than 4 hours prior to culturing.

### Processing of urine specimens

Each urine specimen was utilized for isolation and identification of colonizing bacterial species, urinalysis via 10-parameter urine reagent test strip (LW Scientific, Lawrenceville, Georgia), and generation of urine glycerol stock for long-term storage and re-isolation if needed. Remaining urine was also frozen at -80°C for future analyses. To determine total CFU/ml of urine, each specimen was diluted 1:100, spiral plated on HardyCHROM UTI agar (Hardy Diagnostics, Santa Maria, California) using using an Eddy Jet 2 spiral pater (Neutec Group inc, Farmingdale, NY), and enumerated using a ProtoCOL 3 automated colony counter (Synbiosis). For detection of Gram-positive and Gram-negative bacteria, a 1 µl calibrated inoculating loop (Laboratory Products Sales, Inc, Rochester, NY) was used for semi-quantitative streak-plating on three types of agar from Hardy Diagnostics (Santa Maria, California): Columbia CNA, Bile Esculin (BEA), and MacConkey. All distinct colonies that could be differentiated by morphology, hemolysis, or color were isolated for further analysis.

Gram-positive bacteria from CNA and BEA plates were tested for catalase using 30% hydrogen peroxide and for PYR activity (Hardy Diagnostics). Isolates that were PYR-positive and catalase-negative were suspected to be *Enterococcus* species, and identified to the species level using previously-described primer sets (92). Isolates that were PYR-negative and catalase-positive were suspected to be *Staphylococcus* species, and subjected to Sure-Vue^TM^ SELECT (Fisher Healthcare) to distinguish *Staphylococcus aureus* from coagulase-negative *Staphylococcus* species. PYR-negative and catalase-negative isolates were suspected to be *Streptococcus* species, and were subjected to a Streptex™ Latex Agglutination Test (Thermo Scientific). Suspected *Streptococcus* isolates that did not have a positive Streptex reaction were designated “miscellaneous PYR-negative catalase-negative Gram-positive isolates”.

Gram-negative bacteria from MacConkey plates were identified to the species level whenever possible using API-20E test strips (BioMérieux, Marcy-l’Étoile, France). Isolates identified as *Pseudomonas aeruginosa* via API-20E were confirmed using previously-described primer sets (93). *Proteus mirabilis* and *Proteus vulgaris* isolates were confirmed by swarming motility on blood agar plates (Hardy Diagnostics).

Isolates of a given organism from consecutive urines specimens from the same participant were assumed to be the same strain if they were the same genus and species, the API-20E biotype number varied by no more than two digits (for Gram-negative isolates), and if colony morphology and antimicrobial susceptibility profiles were consistent week-to-week (see below). If a strain was absent in one urine specimen but had been present in the preceding and following specimens from that participant, re-isolation was attempted from the urine glycerol stock. If the strain still could not be detected, it was assumed to be absent from that urine specimen.

### Antimicrobial susceptibility testing

Antimicrobial susceptibility was assessed by zone of growth inhibition on Mueller-Hinton agar (Hardy Diagnostics). Zone diameters indicative of susceptibility were determined using the Clinical and Laboratory Standards Institute (CLSI) breakpoints listed in the M100 30^th^ edition (94, 95). *Enterococcus* isolates were tested for vancomycin sensitivity using Etest strips (Hardy Diagnostics), and a minimum inhibitory concentration of ≤4 ug/ml was considered susceptible. *S. aureus* isolates were tested for methicillin susceptibility using cefoxitin (Hardy Diagnostics), and susceptibility was defined as a zone diameter of ≥18 mm. Gram-negative isolates were tested for susceptibility to ciprofloxacin (Hardy Diagnostics) (≥21 mm zone diameter), ceftazidime (Hardy Diagnostics) (≥18 mm zone diameter), ceftazidime with clavulanate (Hardy Diagnostics) (≥20 mm zone diameter), and imipenem (Hardy Diagnostics) (≥15 mm zone diameter).

### Statistical Analysis

Logistic regression models and Bayesian multilevel longitudinal models were performed using statistical software R v.4.0.1 (R Core Team 2021, https://www.r-project.org/), and JAGS (https://cran.r-project.org/web/packages/rjags/index.html). All logistic regression models were adjusted for participant-level clustering to account for multiple samples per participant. The Bayesian longitudinal models permitted rigorous handling of missing observations in the data.

For longitudinal analysis of occurrences of microbial species (and similarly for signs and symptoms), we considered two Bayesian multilevel models – one for analyzing the *likelihood of occurrence of each individual species* (and similarly individual signs or symptoms), and another for explaining *the total number of microbial occurrences* (and similarly, sign and symptom occurrences). In the following, we describe the two models used for microbial occurrences; analogous models were used for signs and symptoms.

In the first model (model - 1), we quantified the effects of all microbial species, catheter changes, and antibiotic administration on the occurrence of each individual microbial species through a multilevel logistic regression model of the form:

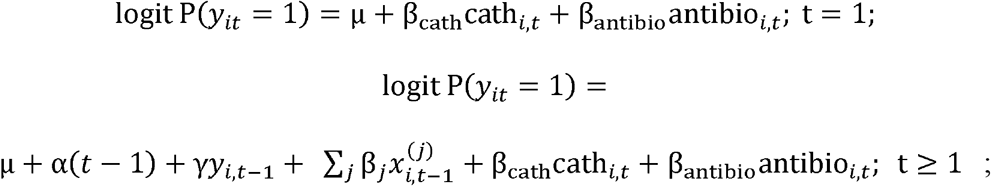

where for an individual *i* at time point *t*, *y_it_* denotes the binary indicator of a specific *response* microbe (e.g. MRSA); 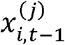 denotes the binary 0-1 indicator of the j-th microbe other than the response microbe at individual *i* at time t-1, and cath*_i,t_* and antibio*_i,t_* denote the binary 0-1 indicators of catheter changes and antibiotic administration, respectively. The parameter *α* quantifies the effect of time, the parameters *γ* and *β_j_*, quantify the logarithms of the odds ratios of the occurrence of the species under consideration, due to one unit change (from 0 (“No”) to 1 (“Yes”); while keeping other predictor variables fixed) in the binary presence status of the same species (e.g., MRSA) and j-th other species *at the previous visit,* and the parameters *β*_cath_ and *β*_antibio_ measure similar log-odds ratios due to one unit change (while keeping other predictor variables fixed) in the binary presence status indicators of catheter change and antibiotic administration *at the current visit* respectively, and for 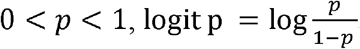. Note that n through the above logistic regression model, missingness in the *response* microbial species occurrence is naturally addressed within a Bayesian statistical framework. To address missingness in the predictor variables, we considered independent Bernoulli distributions

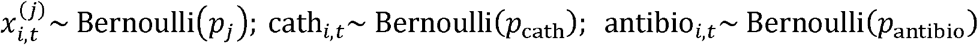

in the second level of the multilevel model. Finally, in the third level of the model we specified (independent, vague) prior distributions for the model parameters. The probability parameters *p_j_*, *p*_cath_, and *p*_antibio_ were assigned independent flat Uniform(0, 1) prior distributions. The intercept μ was assigned an independent Normal(0, 100^2^) prior; the regression parameters *β*_cath_, *β*_antibio_, *α*, and *γ* were assigned independent scale mixture normal priors of the form 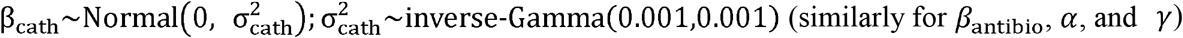. Finally, for the regression coefficients of all ‘other’ microbes, induced shrinkage normal scale mixture priors of the form 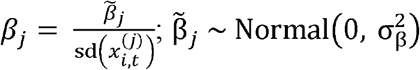; 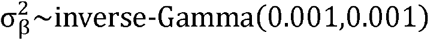 were considered.

In the second model (model – 2), assuming exchangeability of the occurrence of individual microbial species at a given time point in a specific individual, we considered a multilevel Binomial regression model of the form

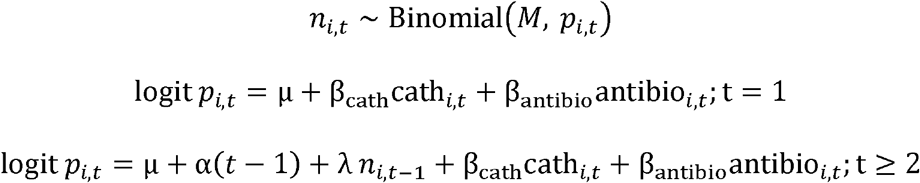

Where *n_i,t_*, is the total number (counts) of observed microbial species, and *p_i,t_*, is the corresponding probability of observing *one generic* microbial species, in subject *i* at time *t*, and *M* denotes the total number of microbial species considered in the study (*M* = 21). Here λ quantifies the log-odds ratio of the occurrence of a *generic* microbial species at time in individual *i* for a one unit increase in the microbial species counts in the same individual at the previous visit, and the remaining parameters have analogous interpretations as in model – 1, with a specific response microbial species being replaced by a *generic* species. Similar to model – 1, missingness in the response *n_i,t_*, are addressed naturally through the model in a Bayesian framework. Missingness in the catheter change and antibiotic administration data are addressed through a similar second (Bernoulli) level:

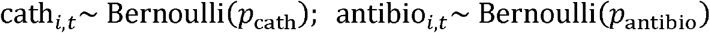

In the final level, independent vague prior distributions similar to model – 1 are considered for the model parameters.

We first fitted model – 1 separately on each response microbial species (and similarly on each response sign or symptom), and then fitted model – 2 collectively on all species (and similarly on all signs and symptoms), by approximating their respective posterior distributions through 10,000 Markov chain Monte Carlo (MCMC) draws, after discarding 10,000 burn-in draws, all generated using JAGS. In a typical Bayesian fashion, missing values in the data were augmented with the respective posterior distributions during the analysis based on the models described above, and were subsequently integrated (marginalized) out. Finally, using the MCMC draws the resulting (marginal) posterior distributions of the various model parameters of interest were summarized through their posterior medians and 95% credible intervals (computed through 0.025^th^ and 0.975^th^ posterior quantiles). These summaries are displayed as forest plots in Supplementary Figures 5 and 6.

## Data Availability

All data are provided within the manuscript and supplemental files.

## Acknowledgments

We would like to thank members of the Department of Microbiology & Immunology, the Division of Infectious Diseases, and Witebsky Center for Microbial Pathogenesis and Immunology for helpful comments and critiques. We would also like to thank the nursing home administrators and staff, as well as the Visiting Nurse Association of Western New York. This work was supported by the National Institutes of Health via the National Institute of Diabetes Digestive and Kidney Diseases [R00 DK105205 and R01 DK123158 to C.E.A.] and the National Center for Advancing Translational Sciences (UL1 TR001412 to the University at Buffalo). The sponsors were not involved in the study design, methods, subject recruitment, data collections, analysis, or preparation of the paper. The content is solely the responsibility of the authors and does not necessarily represent the official views of the funders.

## Author Contributions

Conceptualization, C.E.A.; Methodology, C.E.A. and S.C.; Validation, C.E.A. and A.L.B; Formal Analysis, C.E.A., A.L.B., M.S.H., J.S., and S.C.; Investigation, C.E.A., A.L.B., M.S.H., J.S., and S.C.; Resources, C.E.A.; Data Curation, C.E.A., A.L.B., and M.S.H.; Writing – Original Draft Preparation, C.E.A.; Writing – Review & Editing, all authors.; Visualization, C.E.A. and S.C.; Supervision, C.E.A. and S.C.; Project Administration, C.E.A.; Funding Acquisition, C.E.A..

