## Supplementary figures and images for "Prospective assessment of catheter-associated bacteriuria in nursing home residents: clinical presentation, epidemiology, and colonization dynamics"

### Supplemental Figure 1

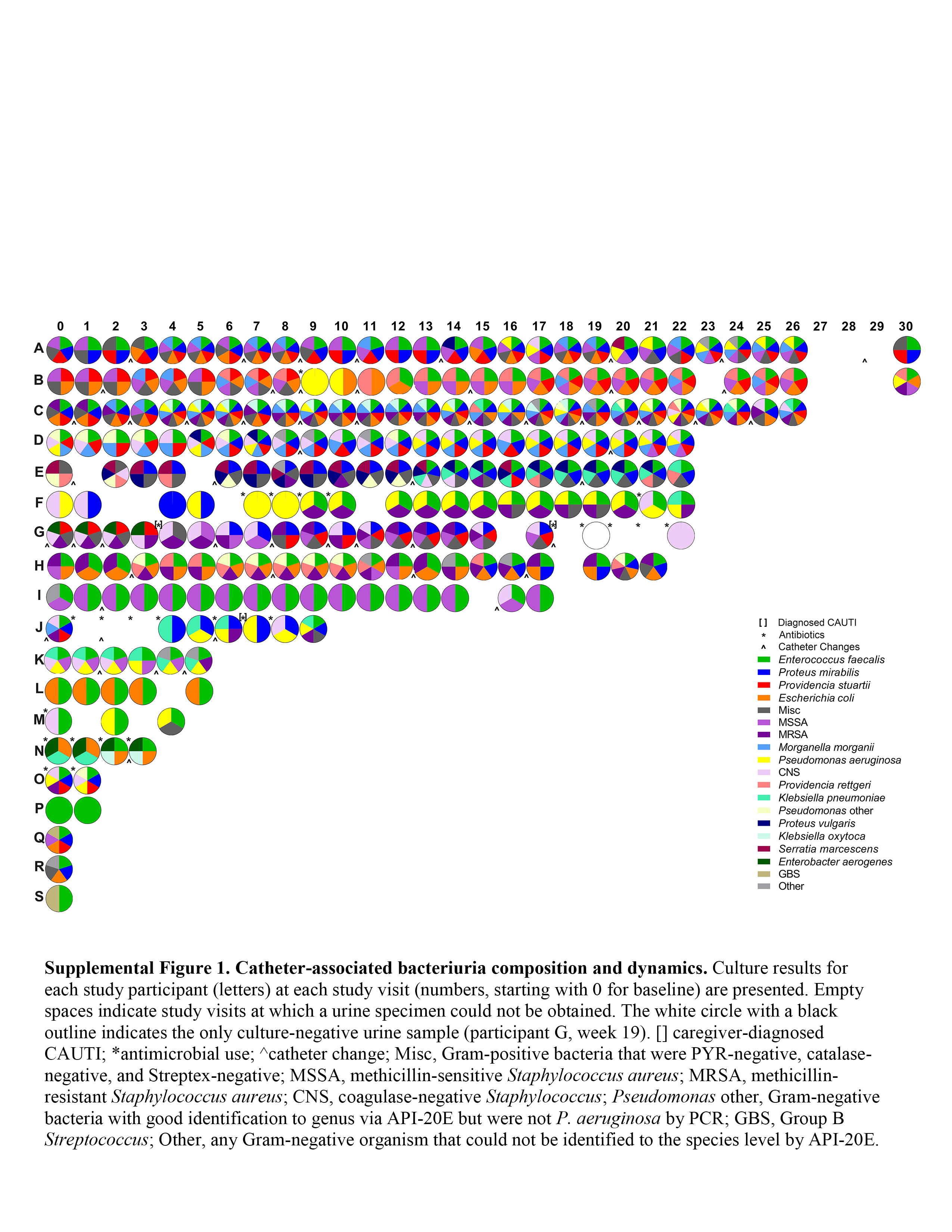

### Supplemental Figure 2

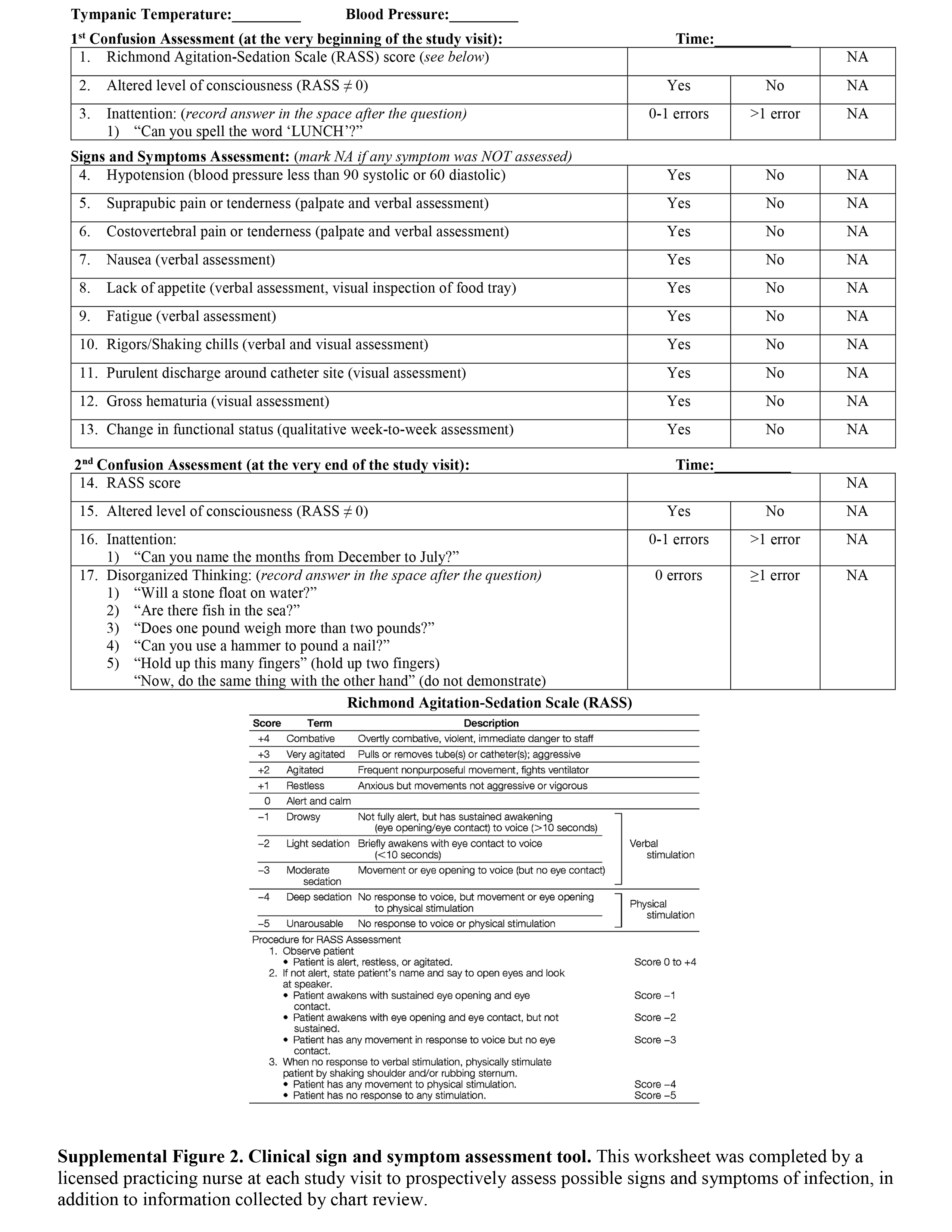

### Supplemental Figure 3

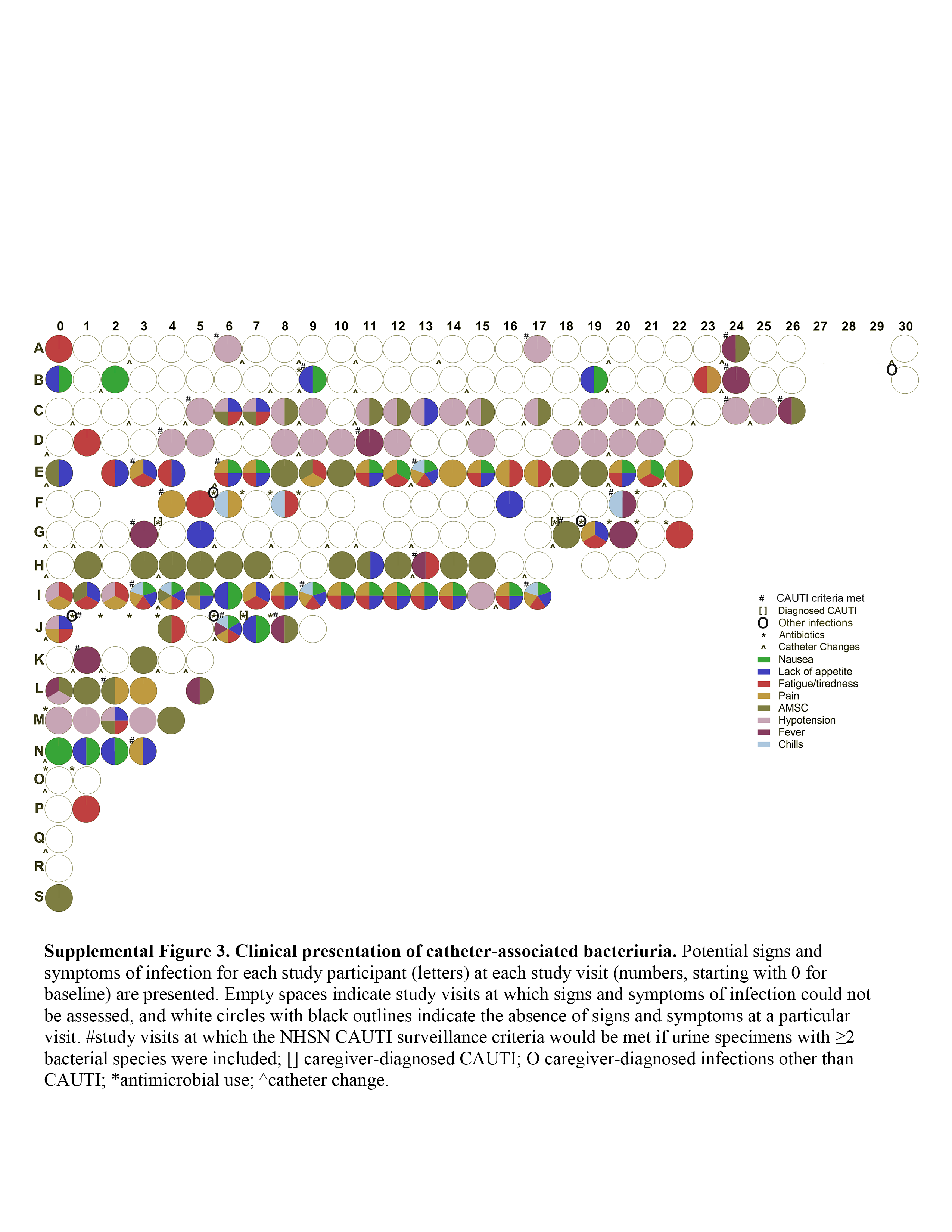

### Supplemental Figure 4

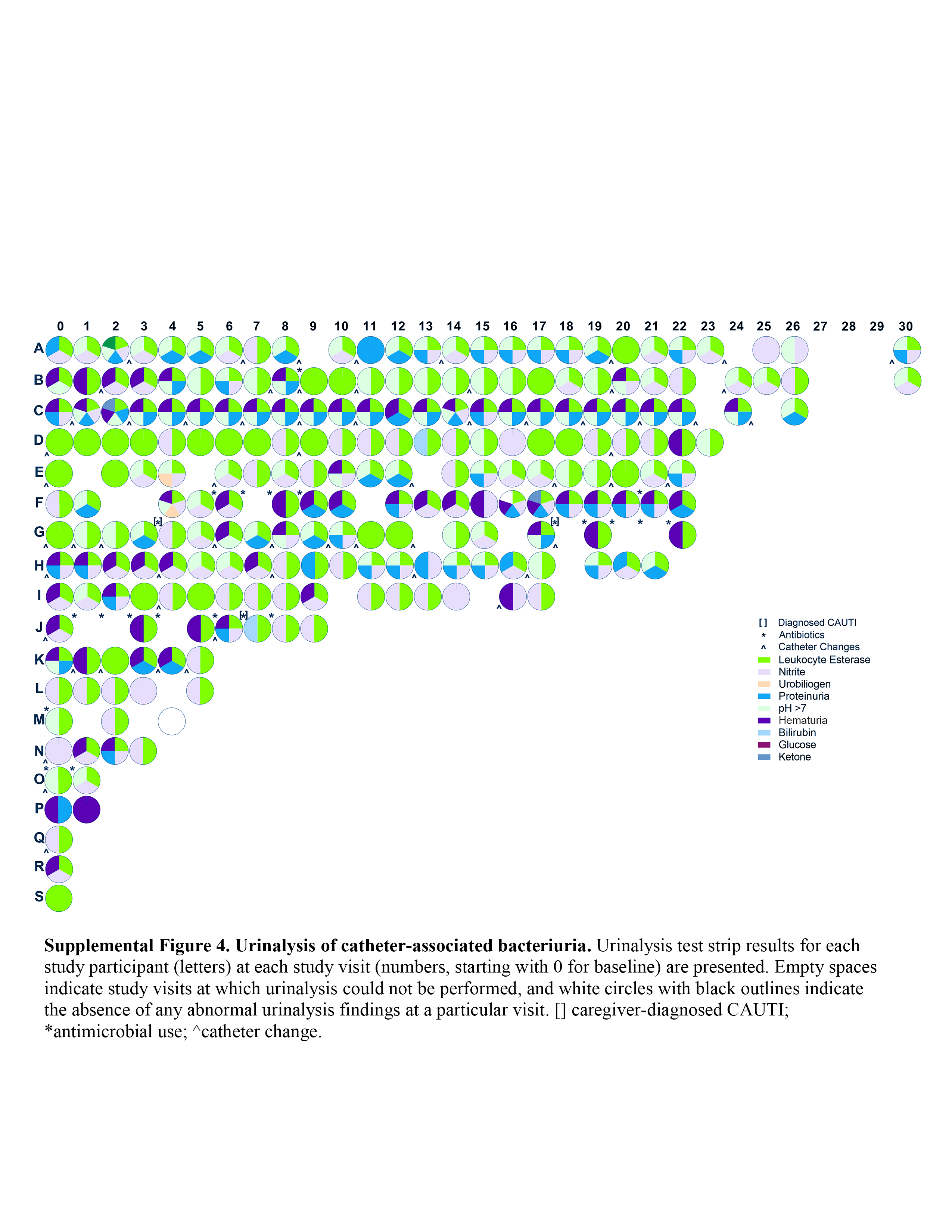

### Supplemental Figure 5

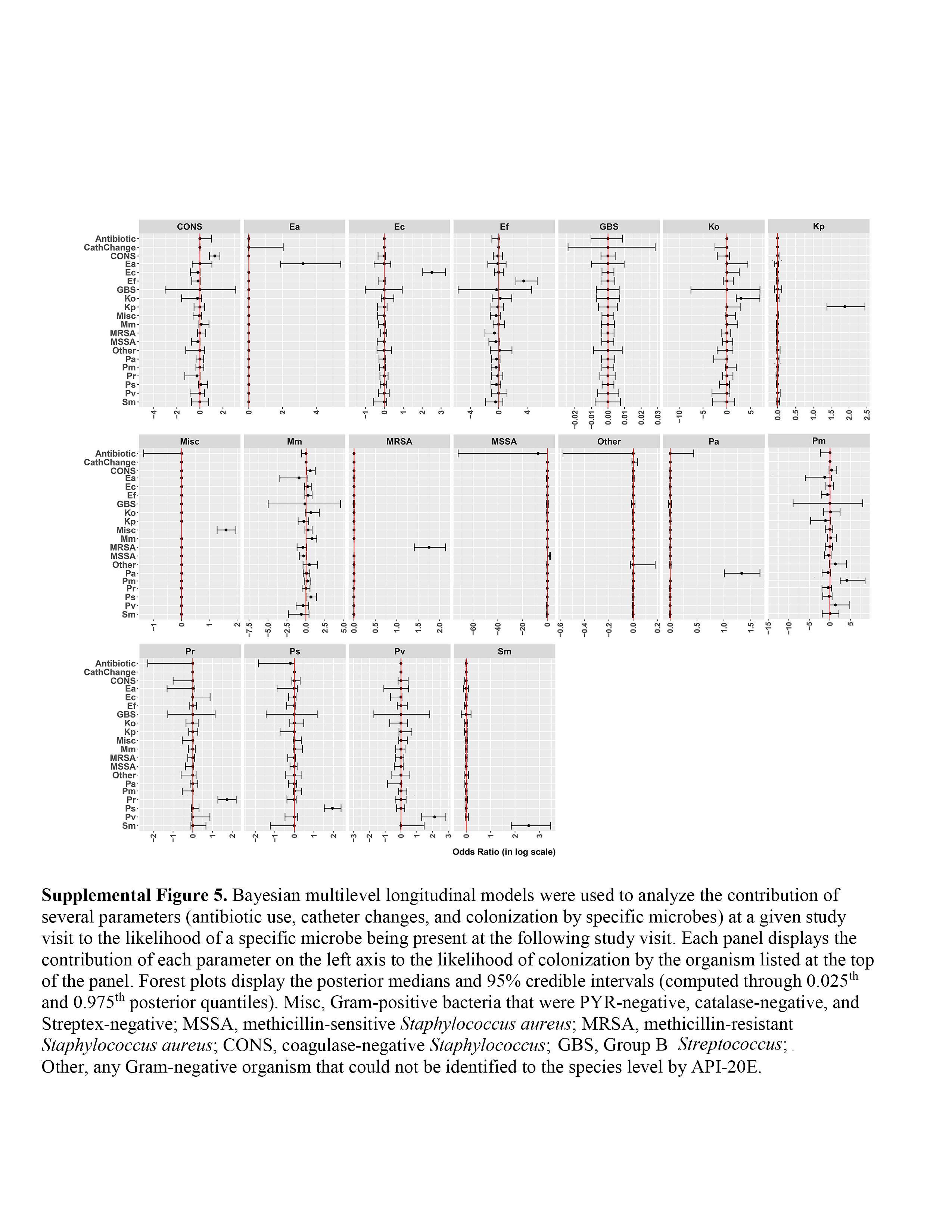

### Supplemental Figure 6

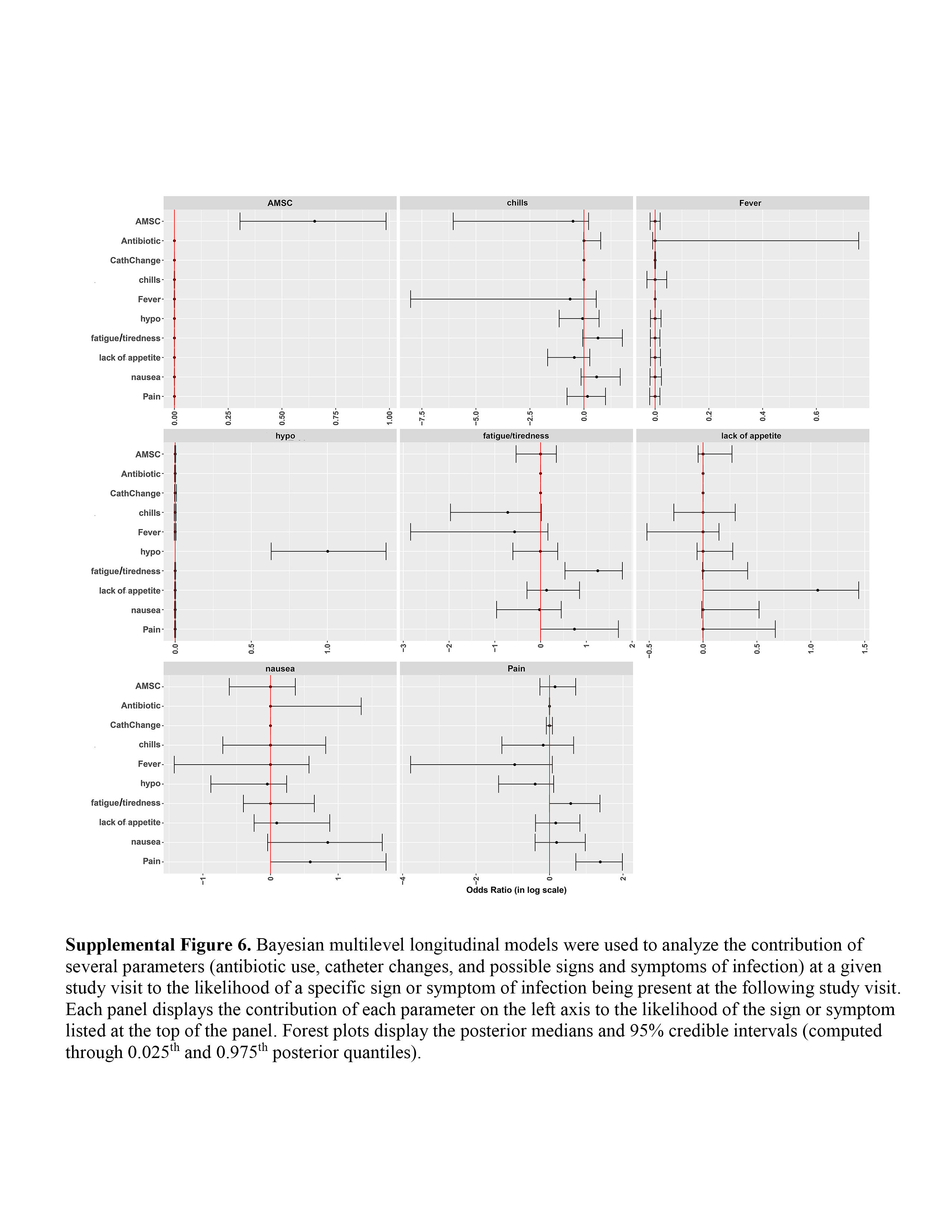
